# Metatranscriptomics-Derived Disease Risk Scores as a Preventive, Diagnostic, and Treatment Support Tool

**DOI:** 10.64898/2026.05.29.26354333

**Authors:** Lan Hu, Michael Bass, Eric Patridge, Matthew Molusky, Grant Antoine, Momchilo Vuyisich, Guruduth Banavar

**Affiliations:** Viome Research Institute, Viome Life Sciences, New York, NY and Bellevue, WA, USA; U.S. Digestive Health Care, PA, USA

**Keywords:** metatranscriptomics, microbiome function, risk stratification, polygenic risk score, preventive health, chronic disease

## Abstract

**Background:** Chronic diseases and symptom syndromes often develop after prolonged biological changes that may precede formal diagnosis. RNA-based metatranscriptomics captures active microbial and human gene expression and may provide a functional layer for disease risk evaluation. To address this translational gap, we developed and validated a Disease Risk Score (DRS) framework that integrates metatranscriptome-derived pathway activity scores from stool, saliva, and blood samples, and evaluated its potential clinical utility as an adjunct risk-evaluation tool.

**Methods:** DRS uses disease-specific sets of pathway activity scores derived from stool and saliva microbial functions, stool and saliva microbial taxa, and blood human gene expression. For each disease, ‘not optimal’ pathway scores are aggregated into a normalized cumulative odds ratio, or cOR, using score-level odds ratios, statistical significance, and literature-supported biological relevance derived from a Development Cohort of 22,369 individuals. A cOR ≥ 5 is defined as high risk. Performance is evaluated in an independent Validation Cohort of 15,908 individuals using self-reported diseases as the reference. Disease support requires both significant cOR separation between self-reported and not-reported (Cohen’s *d* ≥ 0.2) and risk ratio enrichment of self-reported disease among individuals classified as high risk (95% CI of Risk Ratio > 1).

**Results:** Of 20 initially evaluated diseases, 15 meet the prespecified validation criteria on the independent validation cohort: ADHD, anxiety, chronic fatigue syndrome, depression, GERD, hypertension, inflammatory bowel disease, IBS-C, IBS-D, insomnia, MASLD, obesity, obstructive sleep apnea, Sjögren’s syndrome, and type 2 diabetes. Five selected clinical scenarios illustrate how DRS can support clinician-mediated decision making, including IBS subtype reclassification, improved diagnostic acceptance in IBS-D, personalized lifestyle counseling in MASLD and early type 2 diabetes, and diagnostic uncertainty in atypical GERD.

**Conclusions:** DRS is a metatranscriptomics-based risk-stratification framework that aggregates active microbial and human pathway signals into interpretable disease-specific risk estimates across a wide range of disease conditions. Validation against self-reported disease labels in an independent cohort shows significant risk enrichment for each of 15 diseases. DRS is intended as an adjunct to clinical evaluation: a decision support tool in situations where routine care encounters uncertainty, delay, or low patient engagement. Future prospective studies using clinically adjudicated endpoints are needed to assess calibration and clinical outcomes.

## Introduction

Chronic diseases and symptom syndromes often emerge after a prolonged period of biological change, during which inflammatory, metabolic, immune, neurobehavioral, and microbiome activity may shift before a formal diagnosis is made. In routine care, this creates a practical gap between ‘apparently healthy’ and ‘clinically diagnosable’. Patients may have nonspecific symptoms, borderline laboratory results, discordant clinical presentation, or low acceptance of a diagnosis when the biological basis is not apparent. Risk stratification tools can help address this gap by identifying individuals who may benefit from personalized preventive intervention, earlier confirmatory evaluation, or closer monitoring.

Polygenic risk scores (PRSs) provide an important precedent for this type of clinical framework. Rather than relying on a single marker, PRSs aggregate many small, disease-associated genetic effects into a composite risk estimate that can support risk enrichment, screening prioritization, and prevention planning^**1,2**^. Many chronic diseases are influenced by dynamic and modifiable processes, including immune activation, oxidative stress, microbial fermentation, bile acid metabolism, oral and gut barrier function, metabolic regulation, and host inflammatory signaling^**3–9**^. A clinically useful risk framework for these diseases may therefore benefit from integrating functional molecular signals that reflect the individual’s current biological state aligned with such PRS approaches.

RNA-based metatranscriptomics offers such a functional layer. Unlike DNA-based approaches that primarily measure genomic potential, metatranscriptomics measures expressed RNA and therefore captures active microbial and host gene activity at the time of sampling. Stool and saliva metatranscriptomics can quantify both active taxa as well as microbial functions in KEGG Orthologs, while blood transcriptomics can quantify human gene-expression activity. This multi-sample approach is biologically relevant because many chronic diseases involve cross-system interactions among the gut microbiome, oral microbiome, immune system, metabolic tissues, and systemic host responses. Prior multi-omics studies have shown that microbial and host functional patterns are associated with diseases such as inflammatory bowel disease (IBD), irritable bowel syndrome (IBS), and type 2 diabetes (T2D), and prospective studies suggest that microbiome features may precede incident disease in some settings^**10–16**^.

Here, we describe a metatranscriptomics-based Disease Risk Score (DRS) framework designed to align with the logic of PRS while extending it to dynamic, pathway-level molecular activity. DRS integrates disease-associated pathway activity scores derived from an individual’s stool, saliva, and blood samples into disease-specific risk estimates. Stool and saliva features represent microbial functional and taxonomic activity, while blood features represent human gene-expression activity. For each disease, DRS aggregates ‘not optimal’ pathway scores into a normalized cumulative odds ratio (cOR), intended to provide an interpretable estimate of molecular risk burden as a result of gene expression dysregulation.

The goal of DRS is not to diagnose disease or replace established clinical criteria. Rather, DRS is intended as an adjunct risk-evaluation and decision-support tool. In asymptomatic individuals, a high-risk DRS result may support earlier prevention, lifestyle modification, or clinical follow-up. In symptomatic individuals, DRS may help interpret ambiguous presentations, select more targeted next steps, or improve patient understanding and acceptance of a proposed care plan. This clinical utility is particularly relevant in conditions where diagnosis and management are often delayed, uncertain, or dependent on patient adherence, such as IBS subtyping, atypical GERD, MASLD, early T2D, sleep disorders, and inflammatory or immune-mediated conditions.

In this study, we developed the DRS framework (Fig. 1) in a large Development Cohort (N = 22,369) and evaluated its performance in an independent Validation Cohort (N = 15,908) using self-reported diseases as the reference. We assessed both distributional shift of cOR between self-reported and not-reported and enrichment of self-reported disease prevalence among individuals classified as high risk. We further present clinical cases illustrating how DRS may help resolve common decision bottlenecks, including diagnostic reclassification, diagnostic acceptance, targeted treatment selection, and improved adherence to personalized diet and lifestyle recommendations.

**Fig. 1.**
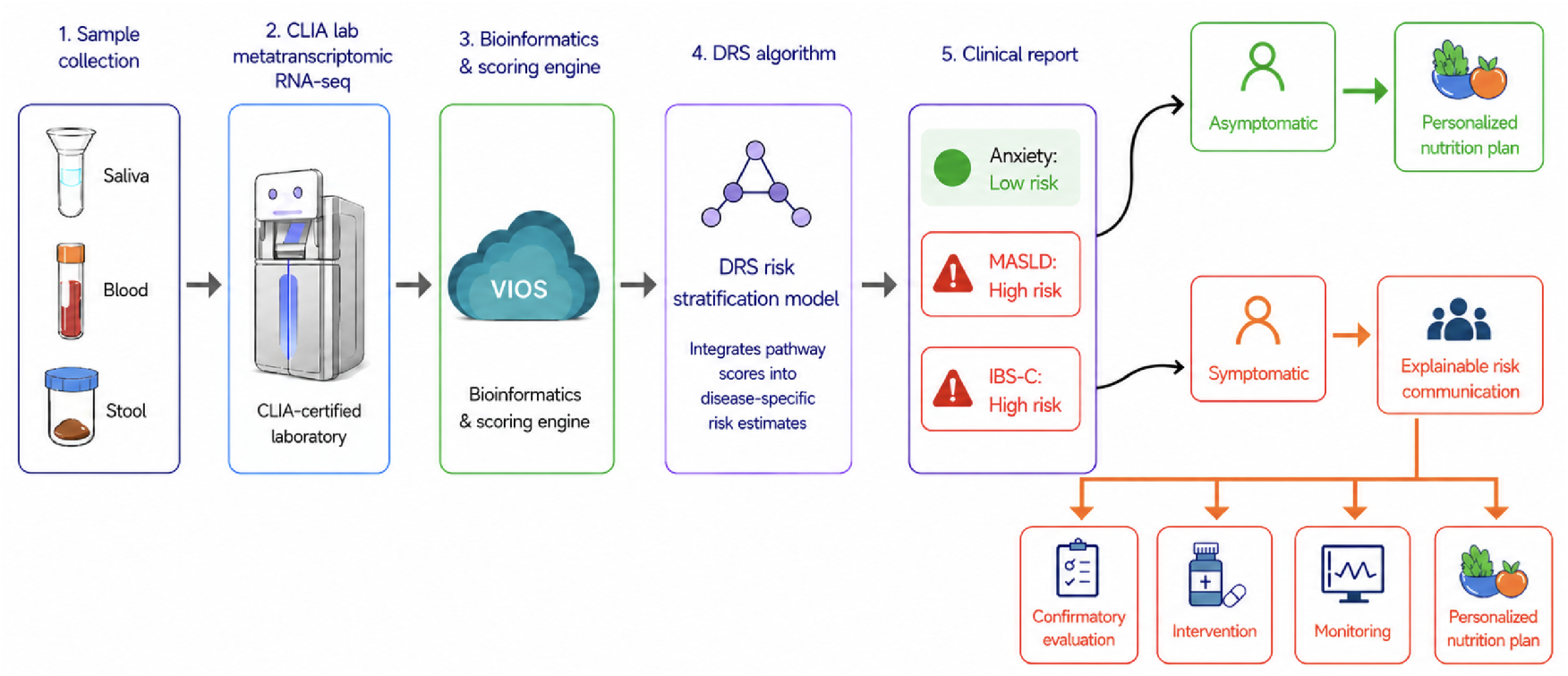
Overview of DRS framework and clinical utility. (1) Stool, saliva, and blood samples are collected for each participant. (2) Samples are sent to Viome’s CLIA certified laboratory for metatranscriptome RNA-sequencing. (3) Bioinformatics pipelines for microbiome (stool and saliva) and human transcriptome (blood) are run in the cloud (ViOS) to quantify the abundance of active microbial functions, taxa, and human genes subsequently. Viome pathway scoring engine then computes the pathway activity scores using these quantification results. (4) The DRS model integrates ‘not optimal’ pathway scores into disease-specific risk estimates. Each disease has its associated pathway scores curated based on literature and Viome’s data. (5) Examples of disease risk levels in the report from the DRS algorithm to facilitate the clinical decision. As illustrated, MASLD (previously known as NAFLD) is ‘high risk’ by DRS, but the patient is asymptomatic, it can prompt personalized dietary and lifestyle modification for prevention. Another example shown is ‘high risk’ IBS-C by DRS and the patient is symptomatic, the specific ‘off’ pathways help with the risk communication for the downstream actions, such as confirmatory evaluation, targeted pharmaceutical intervention, monitoring, and personalized dietary and nutrition plan.

## Methods

### Study population

The source population consists of adults aged 18 years or older who submitted stool, saliva, and blood specimens through Viome’s Full Body Intelligence test and provided authorization for research use of their molecular profiles and questionnaire-derived health information through the terms and conditions completed at enrollment.

Full Body Intelligence kits are shipped directly to participants’ homes with detailed instructions. Each kit includes a stool collection tube, a saliva collection tube, and two blood collection tubes. All tubes are pre-filled with the RNA Preservation Buffer (RPB), which protects RNA integrity from the time of collection and up to 28 days at room temperature^**17–19**^.

After the samples are sent back to Viome, the participants also complete a questionnaire including their demographic, anthropometric measurements, and any diseases or health conditions from a predefined list with over 400 comorbidities (Supplemental Table 1).

We define two mutually exclusive cohorts for development and validation of DRS. The Development Cohort has 22,369 individuals, and is curated based on participant age (≥18 years), BMI (≥14 and ≤60) and their metatranscriptome data quality (see the next section) to represent a robust reference cohort. The Validation Cohort consists of 15,908 individuals with the same age and BMI inclusion criteria. The cohort characteristics are shown in Table 1.

**Table 1.** Cohort characteristics. Age and BMI are mean (SD).

|  | Development | Validation |
| --- | --- | --- |
| N | 22,369 | 15,908 |
| Age | 45.6 (12.9) | 47.4 (13.3) |
| BMI | 25.9 (5.1) | 26.3 (5.4) |
| Female % | 61.5% | 57.1% |

### Metatranscriptomic analysis

RNA molecules from stool, saliva, and blood samples from the participants are extracted and sequenced using previously reported metatranscriptomic techniques^**17–19**^. In the automated lab, total nucleic acids are extracted using magnetic silica beads, and DNA is degraded using a DNase. Ribosomal RNA and a few additional transcripts that are highly abundant are removed from total RNA using a subtractive hybridization method. The remaining transcripts are converted to an Illumina sequencing library using proprietary, dual unique barcodes, and sequenced using the 2×150 bp chemistry.

Active microbial functions in KEGG Orthologs (KO) and species relative abundance are quantified in stool and saliva samples, while human gene expression abundance is quantified in blood samples. See Supplemental Methods for bioinformatics details.

To achieve a robust reference cohort, the samples in the Development Cohort need to meet the predefined sequencing depth criteria. Specifically, stool samples have minimum 500K mapped reads, saliva samples minimum 100K mapped reads, and blood samples minimum 1M mapped reads.

### Pathway scores

A total of 41 pathway scores from stool, saliva, and blood are used in the DRS model (Table 2 and Supplemental Table 2). Each pathway score quantifies the aggregate activity of a set of interacting molecular features (microbial KOs, microbial species, or human genes) relevant to a specific biological or clinical domain. The scores are designed not as diagnostics but as ‘early health insights’ that capture the activity of well-known biochemical processes in health and disease. The development and validation methodology for the stool- and saliva-based pathway scores has been previously described in detail^**20,21**^, and the blood-based scores follow an analogous framework. The scores are developed independently from each other, with key functions, components, metadata, and phenotype labels driving feature selection for each score. The number of features per score ranges from 16 to 63. Across the stool and saliva scores specifically, approximately 93% of KOs appear in only one score, and any two scores share a maximum of three KOs, minimizing redundancy.

**Table 2.** Overview of pathway scores.

| Sample type | Feature type | # scores |
| --- | --- | --- |
| Stool | Microbial KO | 16 |
| Stool | Microbial species | 1 |
| Blood | Human gene | 12 |
| Saliva | Microbial KO | 10 |
| Saliva | Microbial species | 2 |
| **Total** |  | **41** |

The scores have values between 0 and 100, and are standardized to exhibit a Gaussian-like distribution in the population with mean at 50 and standard deviation at 20, higher value representing the ‘good’ direction of the pathway activity. The continuous score value also is categorized into ‘not optimal’ (≤ 35, bottom 25%), ‘good’ (≥ 65, top 25%), and ‘average’ (between 35 and 65, middle 50%) groups.

### DRS development

#### Disease–score association

The foundation of DRS is to establish the relevance of ‘not optimal’ pathway activities to specific chronic diseases quantitatively. Even when individuals may not yet have external symptoms, they are more likely to occur due to an underlying molecular dysfunction identified through ‘not optimal’ pathway scores. A panel of 20 prevalent chronic diseases are selected initially, listed in Table 3. DRS first establishes the association of which disease to which specific Viome’s pathway scores. The curation of such associations is supported by evidence from the extensive scientific literature and Viome’s data analyses. The curation also ranks the associated scores for a disease for their contribution (Supplemental Table 3).

**Table 3.** Cohort characteristics across the initial 20 diseases. Self-report count (prevalence); age and BMI are mean (SD). Dev., Development Cohort (N=22,369); Val., Validation Cohort (N=15,908).

| Disease | Dev. self-report | Dev. Female | Dev. Age | Dev. BMI | Val. self-report | Val. Female | Val. Age | Val. BMI |
| --- | --- | --- | --- | --- | --- | --- | --- | --- |
| Abdominal Weight | 6174 (27.6%) | 72.3% | 47.7 (12.0) | 29.5 (5.5) | 2953 (18.6%) | 67.6% | 49.0 (12.2) | 30.1 (5.9) |
| ADHD | 1433 (6.4%) | 66.2% | 38.9 (10.6) | 26.3 (5.4) | 859 (5.4%) | 67.3% | 40.2 (10.7) | 26.7 (5.9) |
| Anxiety | 4938 (22.1%) | 74.3% | 41.6 (11.7) | 26.2 (5.6) | 2761 (17.4%) | 72.0% | 42.7 (12.2) | 26.4 (5.9) |
| Atherosclerosis | 106 (0.5%) | 41.5% | 64.1 (9.8) | 26.2 (5.3) | 85 (0.5%) | 38.8% | 62.4 (10.2) | 26.9 (5.6) |
| Chronic Fatigue Synd. | 688 (3.1%) | 75.0% | 45.1 (12.7) | 26.2 (5.9) | 456 (2.9%) | 71.7% | 46.5 (13.0) | 26.5 (5.9) |
| Depression | 2446 (10.9%) | 72.3% | 42.5 (12.1) | 27.1 (6.0) | 1414 (8.9%) | 72.2% | 43.9 (12.5) | 27.3 (6.4) |
| GERD | 2943 (13.2%) | 62.9% | 46.8 (12.8) | 27.4 (5.7) | 1863 (11.7%) | 55.2% | 49.2 (13.5) | 27.6 (6.0) |
| Gum Disease | 2293 (10.3%) | 62.7% | 46.6 (12.6) | 26.8 (5.6) | 1625 (10.2%) | 59.4% | 48.7 (12.9) | 27.2 (5.9) |
| Hypercholesterolemia | 1702 (7.6%) | 61.8% | 51.8 (11.9) | 27.4 (5.3) | 1432 (9.0%) | 53.1% | 52.9 (11.8) | 27.5 (5.5) |
| Hypertension | 1792 (8.0%) | 48.9% | 53.6 (12.0) | 29.5 (6.2) | 1485 (9.3%) | 42.9% | 56.2 (11.6) | 29.7 (6.2) |
| IBD | 453 (2.0%) | 65.8% | 44.7 (13.3) | 25.9 (5.8) | 166 (1.0%) | 66.9% | 48.6 (13.9) | 25.8 (5.4) |
| IBS-C | 693 (3.1%) | 84.0% | 45.8 (13.2) | 25.2 (5.3) | 440 (2.8%) | 80.0% | 48.4 (13.8) | 25.1 (5.0) |
| IBS-D | 468 (2.1%) | 63.5% | 45.5 (13.8) | 26.6 (6.1) | 295 (1.9%) | 62.0% | 49.0 (14.6) | 25.8 (5.2) |
| Insomnia | 1153 (5.2%) | 72.9% | 48.7 (13.3) | 26.1 (5.6) | 697 (4.4%) | 70.9% | 51.1 (13.0) | 26.6 (6.0) |
| Kidney Disease | 96 (0.4%) | 56.3% | 56.1 (13.0) | 27.2 (5.4) | 99 (0.6%) | 48.5% | 57.4 (13.6) | 27.3 (5.2) |
| MASLD | 300 (1.3%) | 67.0% | 50.6 (12.0) | 31.1 (6.7) | 218 (1.4%) | 49.5% | 53.1 (12.4) | 31.1 (6.9) |
| Obesity | 1567 (7.0%) | 73.2% | 48.8 (11.8) | 34.6 (5.8) | 818 (5.1%) | 68.0% | 50.7 (12.6) | 35.1 (6.2) |
| Sjögren's Syndrome | 123 (0.6%) | 93.5% | 49.6 (11.6) | 26.5 (6.0) | 111 (0.7%) | 87.4% | 54.2 (12.3) | 25.1 (4.6) |
| Obstructive Sleep Apnea | 1068 (4.8%) | 39.6% | 51.8 (11.9) | 30.6 (6.5) | 810 (5.1%) | 36.5% | 54.1 (12.1) | 30.2 (6.5) |
| T2D | 1641 (7.3%) | 64.5% | 51.6 (12.3) | 29.6 (6.7) | 968 (6.1%) | 53.0% | 55.2 (12.4) | 29.8 (7.0) |

Gum Disease includes both early (gingivitis) and late (periodontitis) gum diseases.

Secondly, in the Development Cohort, respective Case and Control are defined based on the participants’ self-reported diseases or health conditions in the questionnaire. The predefined over 400 comorbidities are binarized for each individual, i.e. 1 as self-reported, 0 as not reported. Participants can select as many as they see fit. For a disease of interest, the Case is defined as having that comorbidity be 1. For the respective Control, not only that particular comorbidity needs to be 0, further detailed curation is performed to exclude likely co-occurring conditions to remove the contamination to our best knowledge given the large list of comorbidities. For example, for IBD, its Case is composed of participants who self-reported IBD; its Control further excludes participants who self-reported colitis, autoimmune gut condition, IBS (any subtype), colon polyps, and/or colorectal cancer in addition to not reporting IBD. This approach makes the risk estimand closer to ‘association with a specific disease’ rather than ‘association with general morbidity’. Full list of comorbidities to exclude for each of the 20 diseases are provided in Supplemental Methods.

Thirdly, for each of the curated disease score pair, (*D, S*), in the Development Cohort, its odds ratio (OR) and p-value are calculated via a logistic regression with the participant’s Case/Control status as the outcome, the score categories ‘not optimal’ and ‘good’ as the variable, and age, sex, BMI as covariates (Supplemental Table 3). The OR *x* is interpreted as: a person with a ‘not optimal’ score *S* is *x* times more likely to report having disease *D* than someone with a ‘good’ score *S*.

#### Disease-level cumulative odds ratio (cOR)

Each disease in DRS has a list of associated pathway scores whose contribution is represented by its OR as described above. We adopt the PRS approach to represent the composite effect from multiple scores to the disease. PRS approaches are widely used in genomics and epidemiology to summarize the cumulative contribution of multiple genetic variants to an individual’s predisposition for a specific disease or trait. In clinical research, PRSs are primarily used to estimate relative genetic risk and support population-level or individual-level risk stratification, rather than to establish a diagnosis or predict disease with certainty.

We define the ‘cumulative odds ratio’ (cOR) to represent the integrated disease level risk assessment from the associated ‘not optimal’ pathway scores that combines the OR of a disease score pair (*D, S*) and its statistical significance of the association in the Development Cohort, as well as the curated rank of the score to the disease. Full details about DRS modeling are provided in Supplemental Methods. The resulting cOR of a disease in an individual is a value between 0 and 10, because values above 10 do not alter the clinical risk category, are often less stable, may overstate interpretable risk, and can create false precision in a screening context. The category of ‘high risk’ in the DRS reporting corresponds to having cOR ≥ 5. This threshold is selected to represent a large-magnitude association rather than a modest elevation in odds. Prior methodological work mapping odds ratios to standardized effect sizes has shown that OR values above approximately 5 correspond to large effect sizes and strong associations^**22**^.

### DRS validation

The Validation Cohort, independent of the Development Cohort and composed of 15,908 individuals, is used to validate the DRS ‘high risk’ (cOR ≥ 5) predictions against their self-reported diseases. While self-reported data serves as the reference for the validation, we acknowledge inherent limitations including recall bias, self-diagnosis accuracy, and the absence of clinical confirmation. These constraints are addressed in the Discussion section.

For each participant, their ‘not optimal’ pathway scores are retrieved. According to the curated disease-score associations in DRS, each disease has its cOR score as the sum of relevant ‘not optimal’ scores’ DRS weights established in the Development Cohort, as well as the classification of ‘high risk’ if cOR is above 5. The diseases to be included for the risk reporting is determined by both cOR distribution separation between self-reported and not-reported groups, as well as the enrichment of disease prevalence in ‘high risk’ group versus the not high risk group in the Validation Cohort.

A two-group t-test is used to compare the distributions of cOR between participants who self-reported the disease and those who did not. Effect size is quantified using Cohen’s *d* (standardized mean difference) to summarize the magnitude of the group difference independent of sample size. Given the expected attenuation of associations due to self-reported outcome misclassification and the tendency for large cohorts to yield statistically significant yet trivial differences, we pre-specified a minimum effect size threshold (Cohen’s *d* ≥ 0.2) to identify diseases exhibiting at least a small, practically meaningful distributional shift.

Using categorical ‘high risk’ prediction, for each disease, we define True positive (TP) as self-reported and predicted high risk, True negative (TN) as not reported and predicted not high risk, False positive (FP) as not reported and predicted high risk, and False negative (FN) as self-reported and predicted not high risk; and p_1_ = P(self-reported=1 | high risk pred=1) = TP/(TP+FP), p_0_ = P(self-reported=1 | high risk pred=0) = FN/ (FN+TN). Then we adopt the risk ratio (RR) as RR = p_1_/p_0_ with its 95% confidence interval (CI) calculated from the standard Katz log method^**23**^ to represent the enrichment of prevalence in high risk vs not high risk^**24**^. The lower 95% CI must be above 1 to be considered acceptable.

## Results

### Cohort characteristics

We selected 20 prevalent chronic diseases as the initial disease panel. They include cardiometabolic (abdominal weight, obesity, MASLD, T2D, hypertension, hypercholesterolemia, atherosclerosis), gastrointestinal (GERD, IBD, IBS-C, IBS-D), neurobehavioral/mental health (ADHD, anxiety, depression), sleep (insomnia, obstructive sleep apnea), oral health (gum disease), renal (kidney disease), and immune system (chronic fatigue syndrome, Sjögren’s syndrome) categories. Table 3 lists the demographic and clinical characteristics in these 20 diseases in the Development and Validation cohorts. The two cohorts exhibit similar breakdowns across the diseases.

### Significantly higher cOR risk score in self-reported cases

Each participant in the Validation Cohort has disease specific cORs computed from their pathways scores (See Methods). Across the initial 20-disease panel, 17 showed a statistically significant separation in cOR between self-reported and not-reported participants (two-group t-test p-value < 0.05). Of these, gum disease and hypercholesterolemia did not meet the prespecified minimum effect size threshold (0.2). Three diseases did not reach statistical significance: abdominal weight, atherosclerosis, and kidney disease. The latter two also had the smallest numbers of self-reported cases (atherosclerosis *n*=85; kidney disease *n*=99), consistent with reduced precision and wider uncertainty intervals.

Figure 2 summarizes the 15 diseases that met both the statistical-significance and effect-size criteria, showing the mean cOR difference (self-reported minus not-reported) with 95% confidence intervals. For all 15 diseases, the estimated mean difference was positive and the lower bound of 95% confidence interval did not cross zero, indicating consistently higher cOR values among self-reported cases in the Validation Cohort.

**Fig. 2.**
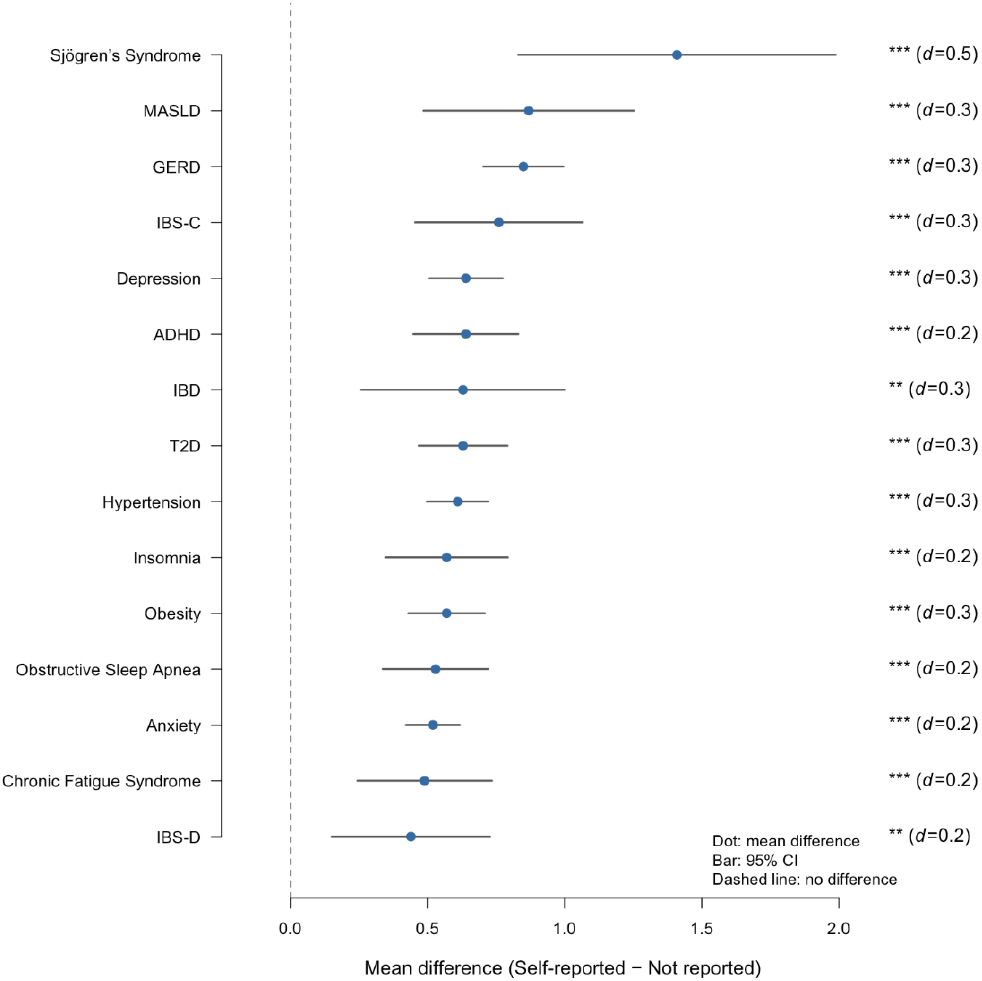
Separation of cOR distributions between self-reported and not-reported of 15 diseases in the Validation Cohort. For each disease, the point denotes the mean difference in cOR (self-reported minus not reported) and the horizontal bar indicates the 95% CI of the difference; the vertical dashed line marks no difference. Only 15 diseases meeting the prespecified criteria for statistical significance in two-group comparisons and minimum effect size are shown. Diseases in the y-axis are ordered by the mean difference. Cohen’s *d* values are shown in the parenthesis to summarize standardized effect size. *: p-value < 0.05, **: p-value < 0.01, ***: p-value < 0.001.

### High risk prediction enriched in self-reported cases

Our DRS method classifies a participant as ‘high risk’ of a disease when its cOR is above 5. We adopt a risk ratio (RR) to compare the disease prevalence in the high risk group versus the not high risk group. For a disease to be qualified for screening in our validation, the lower bound of 95% CI of RR must be above 1.

Figure 3 shows that participants classified as ‘high risk’ by the cOR had a higher prevalence of self-reported disease than those classified as not high risk across all 15 diseases that passed the prespecified screening criteria. For each disease, the estimated RR was >1 and the lower bound of the 95% confidence interval exceeded 1, indicating consistent enrichment of self-reported cases in the high-risk group. The magnitude of enrichment varied by disease, with the largest RRs observed for Sjögren’s syndrome and MASLD, while others showed more modest but statistically supported increases in prevalence.

**Fig. 3.**
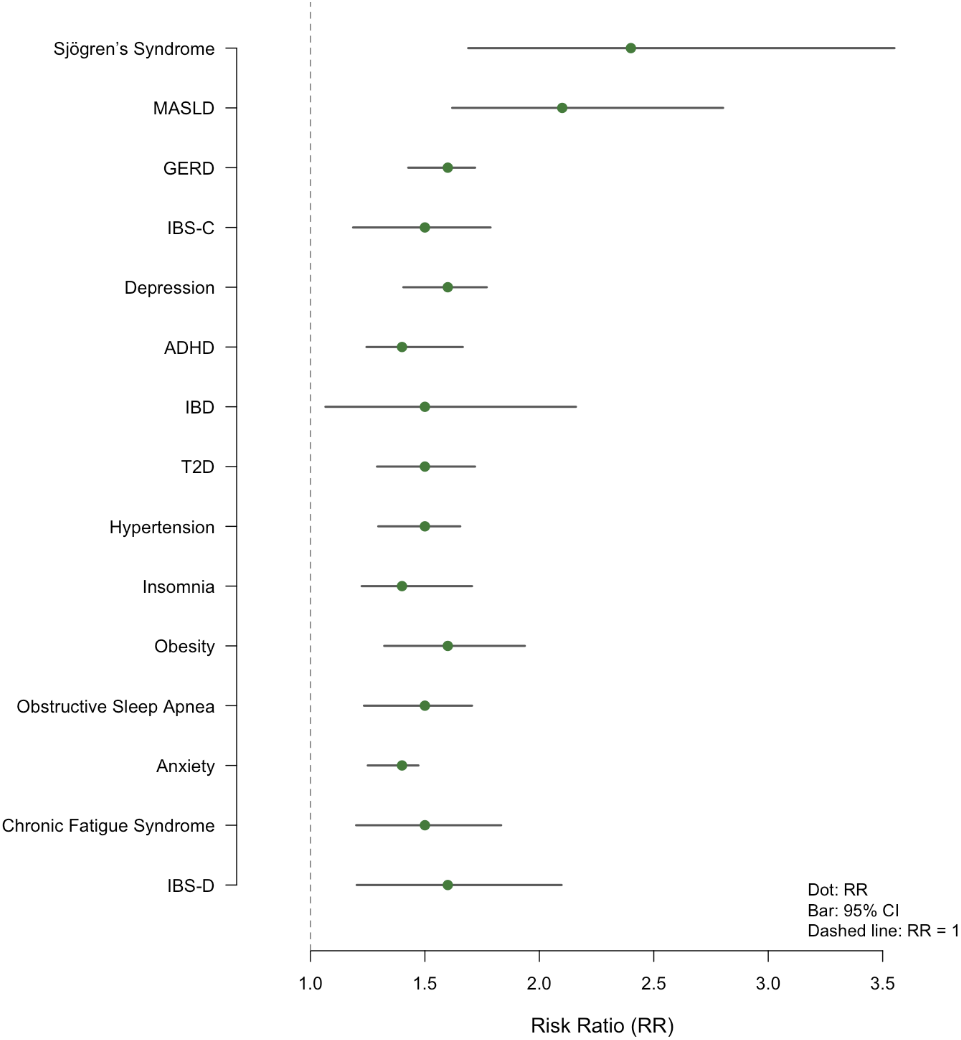
Enrichment of self-reported diseases among participants classified as high risk (cOR ≥ 5) in the Validation Cohort. For each disease, the point denotes RR and horizontal bars indicate 95% CI. The vertical dashed line at RR = 1 marks no enrichment. Diseases in y-axis are shown in the same order as that in Figure 2. The lower bound of the 95% confidence intervals all exceeded 1, indicating consistent enrichment of self-reported cases in the high risk group.

From the initial panel of 20 diseases, 15 have achieved both a statistically significant difference in cOR distributions between self-reported and not-reported in two-group comparisons with minimal 0.2 effect size (Figure 2), as well as accepted ‘high-risk’ classification enrichment (Figure 3) in the Validation Cohort, and thus are selected for DRS risk screening. They are ADHD, anxiety, chronic fatigue syndrome, depression, GERD, hypertension, IBD, IBS-C, IBS-D, insomnia, MASLD, obesity, Sjögren’s syndrome, obstructive sleep apnea, and T2D.

### DRS-based clinical utility

DRS is intended as an adjunct risk screening for providers, supporting earlier and more personalized preventive care, not for disease diagnosis. DRS can provide clinical utility for triage, targeted intervention, patient communication, longitudinal monitoring, and proactive prevention for each of the 15 validated disease areas. As illustrative examples of clinical utility, Table 4 provides five selected clinical scenarios where DRS can improve the patients’ outcomes (the full list of scenarios is too long for a single paper), ranging from diagnostic reclassification, improved diagnostic acceptance and targeted treatment, to improved adherence and lifestyle/diet modifications for early prevention.

**Table 4.** Cases of DRS-informed clinical decisions with improved outcome.

| Case | Disease context | Pre-DRS problem | High-risk DRS | Pathway signal | DRS-informed action | Outcome |
| --- | --- | --- | --- | --- | --- | --- |
| 1 | IBS subtype uncertainty | Apparent IBS-D, treatment failure | IBS-C | Methane gas production 'not optimal' | Imaging for stool burden; shifted to IBS-C therapy | Diagnostic reclassification |
| 2 | Unconvinced IBS-D diagnosis | Low acceptance; trial-and-error care | IBS-D | Sulfide gas production & bile acid metabolism 'not optimal' | Rifaximin; pathway-guided diet | Improved acceptance & targeted treatment |
| 3 | MASLD | Low adherence to lifestyle change | MASLD | Bile acid & oxalate metabolism 'not optimal' | Personalized dietary & lifestyle modification | Improved adherence & acceptance |
| 4 | Borderline T2D | Needs early prevention guidance | T2D | GABA & uric acid production 'not optimal' | Personalized metabolic risk counseling | Earlier prevention planning |
| 5 | Atypical GERD | Diagnostic uncertainty | GERD | Cavity-promoting & oral urease activity 'not optimal' | Trial of GERD medication; tailored diet | Appropriate treatment initiation |

#### Case 1: IBS subtype uncertainty and diagnostic reclassification

The first case involved a patient previously managed as IBS-D because of frequent loose stools, urgency, bloating, and fear of fecal accidents. However, the clinical course was discordant with typical IBS-D management: the patient had not improved with dicyclomine, had difficulty implementing a generic low-FODMAP diet, and continued to report symptoms compatible with stool retention and gas-related distension. A ‘high risk’ IBS-C DRS with a ‘not optimal’ methane gas production pathway reframed the clinical question from ‘persistent IBS-D’ to possible constipation-predominant physiology^**21,25**^ with overflow-type symptoms. This prompted abdominal imaging, which demonstrated significant stool burden, and management was redirected toward IBS-C therapy and methane-informed dietary personalization. The implication is that DRS may help clinicians identify cases where patient-described ‘diarrhea’ reflects overflow physiology rather than primary diarrhea, reducing repeated empiric IBS-D treatment and enabling more appropriate subtype-directed care.

#### Case 2: IBS-D diagnostic acceptance and targeted treatment

The second case involved a patient whose symptoms met an IBS-D pattern, including recurrent cramping, bloating, postprandial urgency, and frequent loose stools, with no alarm features and a prior negative evaluation. The clinical dilemma was not simply diagnostic uncertainty; it was poor diagnostic acceptance. The patient remained concerned that ‘something was being missed’, had failed or discontinued prior empiric therapies, and was requesting repeat imaging and colonoscopy. A ‘high risk’ IBS-D DRS with ‘not optimal’ sulfide gas production and bile acid metabolism pathways made the diagnosis more biologically plausible and actionable^**21,25**^. The sulfide gas production pathway supported a microbiome-active treatment strategy with rifaximin, while the bile acid metabolism pathway supported focused consideration of bile acid mediated diarrhea and pathway guided dietary modification. The implication is that DRS may help move selected IBS-D patients away from repeated exclusionary testing and toward earlier treatment engagement, rational pharmacologic selection, and a more explainable dietary plan.

#### Case 3: MASLD risk communication and lifestyle adherence

The third case involved a patient with hepatic steatosis, mildly elevated liver enzymes, obesity, dyslipidemia, prediabetes, and other cardiometabolic risk factors. Standard evaluation supported MASLD with low initial fibrosis risk, but the clinical dilemma was behavioral: the patient viewed ‘fatty liver’ as a mild incidental finding and found prior advice to ‘lose weight, eat better, and exercise’ too vague to act on. A ‘high risk’ MASLD DRS with ‘not optimal’ bile acid metabolism and oxalate metabolism pathways was used to convert general lifestyle counseling into a more concrete plan. The implication is that DRS may improve MASLD care not by replacing ultrasound, liver enzymes, or fibrosis assessment, but by making early lifestyle intervention more personalized, understandable, and adherable before advanced liver disease develops.

#### Case 4: Early T2D risk and behavioral precision

The fourth case involved a patient with worsening glycemic markers, central adiposity, dyslipidemia, family history of T2D, and lifestyle patterns that included fast food, sugar-sweetened beverages, late-night snacking, and sedentary behavior. Although repeat laboratory testing confirmed early T2D by standard criteria, the patient considered the abnormal values ‘borderline’ and did not feel urgency to change. A ‘high risk’ T2D DRS with ‘not optimal’ uric acid production and GABA production pathways helped make the diagnosis feel more immediate and personally relevant. The uric acid production pathways prioritized elimination of sugar-sweetened beverages, reduction of fructose-heavy and ultra-processed foods, and moderation of alcohol and processed meats. The GABA production pathway provided a biologically plausible link between gut microbial activity and gut-brain signaling, particularly around stress and sleep regulation^**26,27**^. Because sleep disruption and late eating are independently linked to appetite regulation, impaired glucose metabolism, and T2D risk, this pathway helped frame behavioral priorities around stress, sleep timing, and evening meal patterns^**28,29**^. The implication is that DRS may support early T2D intervention by translating generic lifestyle advice into targeted behavioral priorities that patients can understand and follow.

#### Case 5: Atypical GERD and treatment engagement

The fifth case involved a patient with chronic cough, throat clearing, hoarseness, and meal- and position-related symptom worsening, but without classic heartburn or regurgitation. The clinical dilemma was that GERD remained plausible but unproven after unrevealing pulmonary, allergy, and ENT evaluation, while the patient rejected GERD-directed treatment because the diagnosis did not match her understanding of reflux. A ‘high risk’ GERD DRS with ‘not optimal’ oral cariogenic and oral urease activity pathways made the reflux hypothesis more tangible by linking symptoms to oral acid burden and impaired oral pH buffering^**30**^. This supported agreement to a structured, time-limited GERD medication trial, along with tailored dietary and behavioral changes such as reducing acidic beverages, sugar-containing cough drops, grazing, and late-night snacks while supporting oral pH recovery. The implication is that DRS may improve management of atypical GERD by shifting the encounter from diagnostic argument to treatment engagement, while preserving objective reflux testing for nonresponse or recurrence.

### Mechanistic interpretability of selected pathway signals

The cases above demonstrate how DRS results influenced clinical decisions, but the clinical utility of DRS depends on more than disease-level risk categorization. For a DRS result to be useful at the point of care, the accompanying pathway signals must be biologically interpretable. Here we highlight two gas-production pathways that directly influenced the IBS cases: methane gas production in IBS-C subtype reclassification and sulfide gas production in IBS-D treatment selection. The selected pathway examples are intended to demonstrate biological interpretability and decision relevance, not to establish each pathway as an independent diagnostic biomarker.

Methane gas production pathways score is designed around the premise that production of enteric methane slows intestinal transit and augments small intestinal contractile activity, contributing to the constipation phenotype^**31**^. Methane is also naturally correlated with bloating and flatulence, cardinal symptoms of IBS-C.

Sulfide production pathways score is designed around the premise that excessive hydrogen sulfide is cytotoxic: it disrupts colonocyte mitochondrial function, impairs epithelial barrier integrity, and activates inflammatory signaling cascades^**32**^, all mechanisms that contribute to the diarrhea-predominant phenotype.

The inverse relationship between these two pathway signals across IBS subtypes reflects an established competitive dynamic in the gut: sulfate-reducing bacteria and methanogenic archaea compete for overlapping hydrogen and carbon substrates, such that sulfate reduction often restricts methanogenesis. This means that IBS-C individuals, who tend to present with elevated methanogenesis, correspondingly tend to present with lower sulfide production, whereas IBS-D individuals show the opposite pattern^**21,25**^.

## Discussion

### Principal findings

This study describes and evaluates a metatranscriptomics-based Disease Risk Score framework that aggregates disease-associated pathway activity scores from stool, saliva, and blood into disease-specific risk estimates. The framework is conceptually aligned with PRS methods: multiple modest biological signals are combined into a composite score, weighted by association strength, statistical support, and biological relevance. Unlike PRS, however, DRS is based on active microbial and host RNA expression rather than inherited genetic variation, making it a dynamic and potentially modifiable measure of disease-associated biology.

In an independent validation cohort, DRS showed evidence of risk enrichment for 15 of the 20 initially evaluated diseases. For these 15 diseases, self-reported cases had significantly higher cOR values than non-cases, with at least 0.2 standardized effect size in Cohen’s *d*. In addition, participants classified as high risk had a higher prevalence of the corresponding self-reported disease than participants not classified as high risk, with the lower bound of the 95% confidence interval for the risk ratio exceeding 1. These results support the use of DRS as a risk-stratification framework, while also emphasizing that the output should be interpreted as an adjunctive molecular risk signal rather than a diagnostic determination.

The supported 15 diseases span several clinically important domains, including gastrointestinal, cardiometabolic, neurobehavioral, sleep-related, and immune-mediated conditions. This breadth is consistent with the biological design of the framework, which integrates microbial functions, microbial taxa, and human gene-expression activity across multiple sample types. Importantly, DRS does not rely on a single molecular feature or taxon. Instead, it summarizes the cumulative burden of disease-associated ‘not optimal’ pathway signals, which may better reflect the multifactorial biology underlying chronic disease risk.

### Clinical utility: from risk signal to decision support

The clinical value of DRS is best understood not as stand-alone diagnosis, but as decision support in situations where routine care encounters uncertainty, delay, or low patient engagement. The clinical cases illustrate several such use cases. In an IBS subtype case, a high-risk IBS-C DRS result with a ‘not optimal’ methane gas production pathway helped reframe an apparent IBS-D presentation as possible constipation-predominant physiology with overflow symptoms, prompting imaging and subtype-directed management. In an IBS-D case, ‘not optimal’ sulfide gas production and bile acid metabolism pathway signals made the diagnosis more biologically tangible and supported more targeted pharmacologic and dietary intervention. In MASLD and early T2D cases, DRS helped convert generic lifestyle advice into pathway-directed counseling, improving the specificity and perceived relevance of behavior change. In atypical GERD, oral pathway signals supported treatment engagement in a patient whose symptoms did not match their understanding of reflux.

These examples highlight a key distinction: DRS does not replace clinical judgment, guideline-based testing, imaging, laboratory evaluation, or definitive diagnostic criteria. Instead, DRS provides an additional layer of molecular context that can help with the next step by translating a broad risk category into a more actionable care plan by identifying the pathway signals contributing to the risk estimate.

### Metatranscriptomic profiling of dynamic disease-associated biology

RNA-based metatranscriptomics is well suited to risk stratification because it measures expressed RNA and therefore provides a readout of active microbial and host gene expression rather than genomic potential alone. Many disease-associated processes are functional rather than purely taxonomic, including inflammatory tone and host immune signaling^**4,5**^, oxidative stress response^**6**^, microbial gas production^**33**^, bile acid transformation^**34**^, carbohydrate fermentation^**3**^, amino acid metabolism^**35**^, and oral pH regulation^**36,37**^. A DNA-based assay may identify organisms or genes that are present, but RNA expression can provide a closer approximation of what microbial and host systems are doing at the time of sampling.

This distinction is central to the rationale for DRS. Chronic disease risk often reflects a dynamic state shaped by diet, medications, sleep, stress, immune activity, microbiome ecology, and host metabolic regulation. Stress and depression have been linked to gut microbiome, immune, and barrier-related pathways, while chronic systemic inflammation and immunometabolic signaling are recognized contributors to multiple chronic diseases^**4,5,38**^. Prospective studies further support the concept that microbiome features can precede incident disease, including long term prediction of T2D from baseline gut microbiome composition and microbial rhythmicity signatures^**12,13**^. By measuring active pathway expression across stool, saliva, and blood, DRS captures a more current and clinically actionable layer of biology than static inherited risk alone. This also creates an opportunity for longitudinal use: repeated measurement could potentially monitor whether pathway-level risk signals improve, persist, or worsen after diet, lifestyle, pharmacologic, or microbiome-directed interventions.

The multi-sample design is another strength. Stool provides a window into gut microbial function, saliva provides information about oral microbial activity and oral-systemic pathways, and blood provides a host transcriptomic readout that can reflect systemic biology^**8,39–41**^. Many chronic conditions involve interactions across these compartments. For example, cardiometabolic disease may involve gut microbial metabolism, systemic inflammation, and host metabolic signaling; GERD and oral health may involve oral acid burden and microbial urease activity; immune-mediated and gastrointestinal conditions may involve both microbial pathway activity and host inflammatory responses. Integrating these sample types allows DRS to represent disease risk as a systems-level pattern rather than a single-compartment measurement.

### Limitations

Several limitations should be acknowledged. The validation reference is self-reported disease status rather than adjudicated clinical diagnosis. Self-report can introduce recall bias, under-reporting, over-reporting, inaccurate diagnosis labels, and variable disease severity. This limitation may attenuate true associations, but it may also introduce misclassification that cannot be fully resolved analytically. However, the large cohort sizes can nevertheless identify signals from real-world data. Therefore, the current findings support disease risk enrichment against self-reported labels, not diagnostic validity against clinical standards.

The analysis is cross-sectional. As a result, it cannot establish whether DRS signals precede disease onset, reflect existing disease biology, are influenced by treatment, or represent downstream consequences of disease. Medication use, dietary changes, disease duration, and prior interventions may all affect metatranscriptomic activity. For example, lipid-lowering therapy, glucose-lowering therapy, acid suppression, antibiotics, probiotics, or dietary restriction could alter pathway activity and reduce or reshape disease-associated signals. Future analyses should perform longitudinal studies and stratify by treatment status where possible.

Finally, the clinical cases are illustrative rather than definitive evidence of clinical effectiveness. They show plausible use cases in which DRS can contribute to decision making, patient communication, or treatment engagement. However, controlled studies are needed to determine whether DRS-guided care improves outcomes, reduces unnecessary testing, increases adherence, or accelerates appropriate diagnosis and treatment compared with standard care.

### Future directions

Future studies should also evaluate clinical utility directly. Important endpoints include whether DRS changes clinician decision making, improves patient understanding, increases adherence to dietary and lifestyle recommendations, reduces repeated exclusionary testing, or improves symptom and biomarker outcomes. Longitudinal studies can determine whether changes in pathway scores track intervention response and whether DRS can support monitoring over time.

In conclusion, this study supports DRS as a PRS-aligned, metatranscriptomics-based risk stratification framework that identifies enriched risk groups across multiple chronic conditions and provides biologically interpretable pathway signals. The current evidence supports its use as an adjunctive decision-support tool, not as a diagnostic test. Its most promising role is in bridging the gap between molecular risk, clinical ambiguity, and personalized preventive care.

## Data Availability

The datasets used and/or analysed during this study are available from the corresponding author on reasonable request.

## Abbreviations

cOR: cumulative odds ratio
DRS: disease risk score
ENT: ear, nose, and throat
FODMAP: fermentable oligo-, di-, mono-saccharides and polyols
GABA: gamma-aminobutyric acid
GERD: gastroesophageal reflux disease
IBD: inflammatory bowel disease
IBS-C: IBS with constipation
IBS-D: IBS with diarrhea
KO: KEGG ortholog
MASLD: metabolic dysfunction-associated steatotic liver disease
PRS: polygenic risk score
RR: risk ratio
T2D: type 2 diabetes

## Supplementary material

Supplemental Methods.pdf; Supplemental Tables.xlsx.

