## Supplemental Methods for "Metatranscriptomics-Derived Disease Risk Scores as a Preventive, Diagnostic, and Treatment Support Tool"

### Bioinformatics processing

Active microbial functions in KEGG Orthologs (KO) and species relative abundance are quantified in stool and saliva samples, while human gene expression abundance is quantified in blood samples. The microbiome bioinformatics pipeline aligns each sequencing read to microbial genomes as follows: Viome maintains a custom reference catalog which includes 32,599 genomes from NCBI RefSeq release 205 'complete genome' category, 4644 representative human gut genomes of UHGG (1), ribosomal RNA (rRNA) gene sequences, and the human genome GRCh38 (2). These genomes cover archaea, bacteria, fungi, protozoa, phages, viruses, and the human host. The microbial genomes have 98,527,909 total annotated genes. We adopt KEGG Orthology (KO) (3) to annotate the microbial gene functions using eggNOG-mapper (4). The microbiome pipeline maps paired-end reads to this catalog using Centrifuge (5) for taxonomy classification (at all taxonomic ranks). Reads mapped to the host genome and rRNA sequences are tracked for monitoring, but excluded from further analysis. Reads mapped to microbial genomes are processed with an Expectation-Maximization (EM) algorithm (6) to estimate the relative activity of each genome in the sample. Respective taxonomic ranks (strains, species, genera, etc.) can be easily aggregated from the genomes. For this study, we use species relative activity in the downstream analyses. These genome mapped reads are extracted and mapped to only genes or open reading frame (ORF) regions for quantification and KO annotation.

Human transcriptome bioinformatics pipeline uses Salmon (7) to quantify both the transcript and gene expression level. Salmon reference index is created by augmenting the transcripts from Ensemble Release 101 annotations for GRCh38 with 28 selected transcripts from RefSeq. A total of 229,515 transcripts and 60,703 genes are included in the human transcriptome catalog.

4. Huerta-Cepas J, Forslund K, Coelho LP, Szklarczyk D, Jensen LJ, von Mering C, et al. Fast Genome-Wide Functional Annotation through Orthology Assignment by eggNOG-Mapper

[Internet]. 2026 Apr 20 [cited 2026 Apr 20]. Available from: https://dx.doi.org/10.1093/molbev/msx148

5. Kim D, Song L, Breitwieser FP, Salzberg SL. Centrifuge: rapid and sensitive classification of metagenomic sequences. Genome Res. 2016 Dec 1;26(12):1721–9. doi:10.1101/gr.210641.116 PubMed PMID: 27852649.

6. Henderson NC, Varadhan R. Damped Anderson Acceleration With Restarts and Monotonicity Control for Accelerating EM and EM-like Algorithms. J Comput Graph Stat. 2019 Oct 2;28(4):834–46. doi:10.1080/10618600.2019.1594835

7. Patro R, Duggal G, Love MI, Irizarry RA, Kingsford C. Salmon: fast and bias-aware quantification of transcript expression using dual-phase inference. Nat Methods. 2017 Apr;14(4):417–9. doi:10.1038/nmeth.4197 PubMed PMID: 28263959; PubMed Central PMCID: PMC5600148.

### Control comorbidity exclusion sets

**Definition:** for condition **X**, its **controls** are people with X=0 AND **exclude** anyone who has *any* comorbidity in the condition's exclusion set below.

**1) Abdominal Weight**

Exclude controls with:

Obese / Obesity, Metabolic Syndrome, Insulin Resistance, Prediabetes, Abnormal Fasting Glucose, Dysglycemia / Irregular Blood Sugar Type 2 Diabetes, Diabetes, Nonalcoholic Fatty Liver Disease (NAFLD) / SteatoHepatitis (NASH), Sleep Apnea, Hypertension / High Blood Pressure, Hyperlipidemia / High Lipids, Hypercholesterolemia / High Cholesterol, Hypertriglyceridemia / High Triglycerides

**2) ADHD**

Exclude controls with:

Attention-Deficit Disorder (ADD), Autism / Autism-Spectrum Disorder, Asperger Syndrome / Asperger's, Dyslexia Anxiety, Depression, Behavioral / Mood Disorder

**3) Anxiety**

Exclude controls with:

Panic Attack, Post-Traumatic Stress Disorder (PTSD), Obsessive-Compulsive Disorder (OCD), Phobia(s) Depression, Major Depressive Disorder, Bipolar Disorder, Insomnia, Substance Addiction, Alcohol Addiction, Nicotine Addiction, Recreational Drug Addiction, Dysglycemia / Irregular Blood Sugar, Hypoglycemia / Low Blood Sugar, Abnormal Fasting Glucose, Diabetes Gestational Diabetes, Latent Autoimmune Diabetes in Adults (LADA), Prediabetes Type 1 Diabetes, Type 2 Diabetes

**4) Atherosclerosis**

Exclude controls with:

Intracranial atherosclerosis, Coronary heart disease, Peripheral artery disease, Angina, Coronary Microvascular Disease, Myocardial Infarction (MI) / Heart Attack, Congestive Heart Failure, Heart failure Stroke, Transient Ischemic Attack (TIA), Cerebrovascular disease, Hypercholesterolemia / High Cholesterol, Hyperlipidemia / High Lipids, Hypertension / High Blood Pressure, Type 2 Diabetes, Diabetes, Metabolic Syndrome, Renal artery stenosis, Kidney Disease, Hypertriglyceridemia / High Triglycerides

**5) Chronic Fatigue Syndrome**

Exclude controls with:

Fibromyalgia, Chronic Pain Brain Fog, Poor Memory, Mild Cognitive Impairment, Cognitive Dysfunction / Cognitive Impairment, Insomnia, Depression, Anxiety, Major Depressive Disorder, Lyme Disease, Epstein-Barr Virus Infection (EBV), Hepatitis C Virus (HBC), Hepatitis B Virus (HBV), Hepatitis A, Hepatitis, Hypothyroid / Hypothyroidism

**6) Depression**

Exclude controls with:

Major Depressive Disorder, Seasonal Affective Disorder (SAD), Anxiety, Panic Attack, Post-Traumatic Stress Disorder (PTSD), Obsessive-Compulsive Disorder (OCD), Bipolar Disorder, Insomnia, Chronic Fatigue Syndrome, Fibromyalgia, Chronic Pain, Substance Addiction, Alcohol Addiction

**7) GERD**

Exclude controls with:

Barrett's Esophagus, Hiatal Hernia, Eosinophilic Esophagitis, Dyspepsia / Indigestion, Gastritis, Peptic Ulcer Disease, Helicobacter Pylori Infection, Achlorhydria / Hypochlorhydria, Gastroparesis

**8) Gum Disease**

Exclude controls with:

Bleeding Gums, Receding Gums (Gingival recession), Recurring Cavities (Tooth Decay), Tartar (Dental Calculus), Toothache (Dental Abscess), Periodontal cyst (epithelium-lined sac containing fluid), Peri-Implantitis (Inflammation surrounding dental implants), Salivary Gland Infection (Sialadenitis), Swelling or Irritation of the Mouth (Mucositis), Swollen Mucus Membrane (Lichen Planus)

**9) Hypercholesterolemia**

Exclude controls with:

Hyperlipidemia / High Lipids, Hypertriglyceridemia / High Triglycerides, Atherosclerosis, Intracranial atherosclerosis, Coronary heart disease, Angina, Coronary Microvascular Disease, Peripheral artery disease,Transient Ischemic Attack (TIA), Cerebrovascular disease, Hypertension / High Blood Pressure, Type 2 Diabetes, Diabetes, Metabolic Syndrome, Obese / Obesity, Abdominal Weight Gain / Overweight, Stroke

**10) Hypertension**

Exclude controls with:

Cardiovascular Condition / Heart Disease, Atherosclerosis, Intracranial atherosclerosis, Coronary heart disease, Angina, Coronary Microvascular Disease, Myocardial Infarction (MI) / Heart Attack, Congestive Heart Failure, Heart failure, Atrial Fibrillation, Valvular Heart Disease, Cardiomyopathy, Peripheral artery disease, Stroke, Transient Ischemic Attack (TIA), Cerebrovascular disease, Renal artery stenosis, Kidney Disease, Hyperlipidemia / High Lipids, Hypercholesterolemia / High Cholesterol, Hypertriglyceridemia / High Triglycerides, Metabolic Syndrome, Obese / Obesity, Abdominal Weight Gain / Overweight, Type 2 Diabetes, Diabetes, Nonalcoholic Fatty Liver Disease (NAFLD) / SteatoHepatitis (NASH)

**11) IBD**

Exclude controls with:

Microscopic Colitis, Colitis Proctitis (Inflammation of the rectum lining), Anal Fistula, Ischemic Colitis, Autoimmune Gut Condition, Irritable Bowel Syndrome (IBS), Irritable Bowel Syndrome - Diarrhea (IBS D), Irritable Bowel Syndrome - Mixed Type (IBS M), Irritable Bowel Syndrome - Constipation (IBS C), Colorectal Cancer, Benign Tumor / Non-Cancerous Polyps, Colon Polyps

**12) IBS-C**

Exclude controls with:

Irritable Bowel Syndrome (IBS), Irritable Bowel Syndrome - Diarrhea (IBS D), Irritable Bowel Syndrome - Mixed Type (IBS M), Small Intestinal Bacterial Overgrowth (SIBO), Dysmotility, Leaky Gut / Intestinal Hyperpermeability, Dysbiosis, GI Inflammation / Gut Inflammation, Dyspepsia / Indigestion, Gastritis, Acid Reflux / Gastroesophageal Reflux Disease (GERD), Celiac Disease, Diverticulitis, Diverticulosis, Diverticular Condition, Diverticular Condition

**13) IBS-D**

Exclude controls with:

Irritable Bowel Syndrome (IBS), Irritable Bowel Syndrome - Constipation (IBS C), Irritable Bowel Syndrome - Mixed Type (IBS M), Small Intestinal Bacterial Overgrowth (SIBO), Dysmotility, Leaky Gut / Intestinal Hyperpermeability, Dysbiosis, GI Inflammation / Gut Inflammation, Dyspepsia / Indigestion, Gastritis, Acid Reflux / Gastroesophageal Reflux Disease (GERD), Celiac Disease, Diverticulitis, Diverticulosis, Diverticular Condition, Diverticular Condition

**14) Insomnia**

Exclude controls with:

Sleep Apnea, Restless Leg Syndrome, Anxiety, Depression, Major Depressive Disorder, Post-Traumatic Stress Disorder (PTSD), Chronic Pain, Fibromyalgia

**15) Kidney Disease**

Exclude controls with:

Kidney Stones, Kidney Cyst(s), Renal artery stenosis, Renal Cancer / Kidney Cancer, Hypertension / High Blood Pressure, Type 2 Diabetes, Diabetes, Prediabetes, Abnormal Fasting Glucose, Iron-Deficiency Anemia

**16) MASLD/NAFLD**

Exclude controls with:

Liver Condition, Hepatitis,Hepatitis A, Cirrhosis / Liver Cirrhosis, Liver cyst(s), Liver Cancer, Alcohol Addiction, Gallstones, Cholecystitis (Inflamed Gallbladder), Cholestasis (Impaired Bile Flow), Sclerosing Cholangitis, Biliary Insufficiency, Metabolic Syndrome, Insulin Resistance, Prediabetes, Type 2 Diabetes, Diabetes, Obese / Obesity, Abdominal Weight Gain / Overweight, Sleep Apnea, Hypertension / High Blood Pressure, Hyperlipidemia / High Lipids, Hypercholesterolemia / High Cholesterol

**17) Obesity**

Exclude controls with:

Abdominal Weight Gain / Overweight, Metabolic Syndrome, Insulin Resistance, Prediabetes, Abnormal Fasting Glucose, Dysglycemia / Irregular Blood Sugar, Type 2 Diabetes, Diabetes Nonalcoholic Fatty Liver Disease (NAFLD) / SteatoHepatitis (NASH), Sleep Apnea, Hypertension / High Blood Pressure, Hyperlipidemia / High Lipids, Hypercholesterolemia / High Cholesterol, Hypertriglyceridemia / High Triglycerides

**18) Sjögren’s Syndrome**

Exclude controls with:

Autoimmune Disease, Lupus, Rheumatoid Arthritis (RA), Vasculitis Scleroderma, Connective Tissue Disease, Mixed Connective Tissue Disease, Psoriatic Arthritis, Ankylosing Spondylitis, Axial Spondyloarthritis, Dermatomyositis, Polymyositis, Autoimmune Myopathy, Autoimmune Skin Condition, Dry Eye, Dry Mouth (Xerostomia), Autoimmune Joint Condition, Hashimoto's Disease (Autoimmune Thyroid Condition), Hashimoto's Disease (Autoimmune Thyroid Condition)

**19) Sleep Apnea**

Exclude controls with:

Obese / Obesity, Abdominal Weight Gain / Overweight, Metabolic Syndrome, Hypertension / High Blood Pressure, Type 2 Diabetes, Diabetes, Insulin Resistance, Cardiovascular Condition / Heart Disease, Coronary heart disease, Angina, Heart failure, Congestive Heart Failure, Atrial Fibrillation Respiratory Condition, Chronic Obstructive Pulmonary Disease (COPD), Asthma,Insomnia

**20) T2D**

Exclude controls with:

Abnormal Fasting Glucose, Dysglycemia / Irregular Blood Sugar, Insulin Resistance, Metabolic Syndrome, Type 1 Diabetes, Gestational Diabetes, Latent Autoimmune Diabetes in Adults (LADA), Obese / Obesity, Abdominal Weight Gain / Overweight, Nonalcoholic Fatty Liver Disease (NAFLD) / SteatoHepatitis (NASH), Sleep Apnea, Hypertension / High Blood Pressure, Hyperlipidemia / High Lipids, Hypercholesterolemia / High Cholesterol, Hypertriglyceridemia / High Triglycerides, Coronary heart disease, Atherosclerosis, Intracranial atherosclerosis, Heart failure, Stroke, Peripheral artery disease, Retinopathy, Kidney Disease

### Disease level cOR algorithm

1. In the Development Cohort, for each disease-associated score, its overall risk index, a value between 0 and 1, is calculated as a weighted sum from normalized OR, association significance derived from OR p-value, and priority using

$$\textbf{Risk Index} = w_1 \cdot \textbf{OR\_norm} + w_2 \cdot \textbf{Sig\_score} + w_3 \cdot \textbf{Rank\_score}$$

where:

- `OR_norm = (OR - 1) / (max(OR) - 1)` to normalize the effect size (negative if OR<1)
- $\texttt{Sig\_score} = -\log_{10}(\texttt{p-value}) / \max(-\log_{10}(\texttt{p-value}))$ to transform p-values to significance strength (p-values are capped at 0.001)
- `Rank_score = (num_score + 1 - priority_rank) / num_score` to convert priority rank to a linear scale, the higher the better
- $w_1$, $w_2$, $w_3$ are weights summing to 1: $w_1=0.5$, $w_2=0.3$, $w_3=0.2$ to give primary importance to OR effect size, moderate importance to statistical significance, and some consideration to the priority ranking.

2. To support explainability and alignment with epidemiology practice, each disease's combined risk index from associated scores is scaled to 10. The resulting scaled risk index for each score is thus its contributing DRS weights (see Supplemental Table 3).
3. The cOR of a disease in an individual is the sum of DRS weights across the 'not optimal' associated scores between 0 and 10. The category of 'high risk' in our disease risk stratification reporting corresponds to having cOR ≥ 5.
